# Red flags useful to screen for gastrointestinal and hepatic diseases in patients with shoulder pain: a scoping review protocol

**DOI:** 10.1101/2021.03.23.21254088

**Authors:** A Roncone, F Fiorentino, D Pennella, F Maselli, F Brindisino, S Giagio

**Author notes:** **CORRESPONDING AUTHOR:** Fabrizio Brindisino, via della Libertà, 14 73023, Lizzanello (Le), ITALY, mail.

## Abstract

Shoulder pain (SP) is the third most common musculoskeletal disorder worldwide and its prevalence ranges from 5 to 47%.

However, the clinical presentation of signs and symptoms may hide other serious conditions; the so-called Red Flags (RFs).

For these reasons, clinicians dealing with neuro-musculoskeletal shoulder disorders should pay particular attention during the medical history and clinical examination screening and identifying signs that may cause probable referred symptoms.

Considering the variety of clinical presentations of SP and the importance of the screening for referral, identifying and summarizing the possible gastrointestinal and hepatic diseases among these patients may give a comprehensive overview to the clinicians and consequently may improve the overall management.

To the authors knowledge, there are no published studies on the topic and, in this context, a scoping review is strongly required and corresponded to the objectives of this project.

## BACKGROUND

Shoulder pain (SP) is the third most common musculoskeletal disorder worldwide [1], and its prevalence ranges from 5 to 47% [2].

In most cases, SP has neuromusculoskeletal causes such as rotator cuff tear [3], rotator cuff tendinopathy [4], labral tear [5], bursitis [6] and long head of biceps tendinopathy [7].

However, the clinical presentation of signs and symptoms is not always clear and may hide other serious conditions [8]; the so-called Red Flags (RFs) [9][10].

Besides, patients do not directly associate these symptoms, omitting to refer them to the physical therapist (PT) [11].

For these reasons, clinicians dealing with neuro-musculoskeletal shoulder disorders should pay particular attention during the medical history and clinical examination screening and identifying signs that may cause probable referred symptoms [12]. For example, some gastrointestinal and hepatic diseases report pain patterns that may seem musculoskeletal disorders affecting the upper quadrant [12].

Although these competences should be an essential requirement in the daily clinical practice [8], some authors highlighted low screening rates among PTs [10] and general practitioners during the first assessment of the patients [9].

To the authors ‘ knowledge, there are no published studies on the topic.

In this context, a scoping review is strongly required and corresponded to the objectives of this project.

In particular, the present scoping review will aim to:

a. Systematically map and summarize the current literature to identify any studies that reported RFs for gastrointestinal and hepatic diseases among patients who reported SP;
b. Identify gaps in the evidence base and direct future research in this area.

### Review question

The following research question was formulated: “What is known from the existing literature about the RFs for gastrointestinal and hepatic diseases in patients with SP?”

## METHODS

This scoping review will be conducted in accordance with the latest review process proposed by the Joanna Briggs Institute (JBI) in 2020 [13].

For the reporting, the Preferred Reporting Items for Systematic Reviews and MetaLAnalyses extension for Scoping Reviews (PRISMALScR) [14] checklist will be used.

### Inclusion criteria

Full-text articles will be eligible for inclusion if they meet the following population, concept, and context (PCC) criteria:

- Population. This review will consider studies that included patients of any age and gender with any diagnosis of SP.
- Concept. This review will consider studies that explored and reported RFs for gastrointestinal and hepatic diseases among the population described above.
- Context. This review will consider studies conducted in any context Sources. This scoping review will consider any study designs or publication type for inclusion. No time, geographical, setting and language restrictions will be applied.

### Exclusion criteria

Studies that do not meet the above-stated PCC criteria will be excluded.

### Search strategy

Literature research will be carried out on the following databases up to February, 21^st^ 2021: MEDLINE, Web of Science, Cochrane Library and SciELO. The full search strategy for MEDLINE is available in the **Appendix 1**. Additional records will be identified through grey literature (e.g. Google Scholar). We will check the reference lists of all identified studies.

### Study selection

Search results will be collated and imported to EndNote V.X9 (Clarivate Analytics, PA, USA). Duplicates will be automatically removed. The review process will be carried out by two independent authors and it will consist of two levels of screening using Rayyan QCRI online software [15]: (1) a title and abstract review and (2) a full-text review. If an agreement cannot be reached, this will be resolved by means of consolidation with an independent third reviewer. Reasons for the exclusion of any full-text source of evidence will be recorded and reported in the scoping review report. The results of the search will be reported in full in the final scoping review and presented in a Preferred Reporting Items for Systematic Reviews and Meta-analyses (PRISMA) flow diagram.

### Data extraction

To sort out the included studies and extracted data, a standardized planned Excel form will be used. This extraction form will be developed in line with the PCC model and it will be filled in by two authors alternately, with mutual check on each entry. A draft of the extraction tool is provided in the **Appendix 2**.

Charting results is commonly an iterative process during scoping reviews; other data can be added to this form according to the subgroups that could emerge from the analysis of the included studies. Modifications will be detailed in the full scoping review.

### Data synthesis

As a scoping review, the purpose of this study is to aggregate the findings and present an overview on the research rather than to evaluate the quality of the individual studies [13]. The results will be presented in two ways.

1. Numerically: We will summarize and report collated data as a descriptive analysis: mapping the data, showing the distribution of studies by a period of publication, study design, and theme. Results will be reported in tabular and diagrammatic form.

2. Thematically: A thematic summary pertaining to gastrointestinal and hepatic RFs will be performed. Additional descriptive subgroups analysis will be reported (e.g. gender of the patients, shoulder disorders).

## RELEVANCE AND DISSEMINATION

The findings of this scoping review may increase clinicians ‘ knowledge about the identification of RFs in patients with SP and consequently, may suggest a prompt referral. On the whole, this review may add relevant information contributing to the improvement of the clinical management.

The results will be published in a peer-review journal and will be presented at relevant national and international scientific events.

## Data Availability

all data referred to manuscript will be pubblished

## AUTHORS ‘ CONTRIBUTIONS

All authors conceived, designed, drafted and approved the final protocol.

## APPENDIX

**Appendix 1**. Search strategy for MEDLINE.

**Appendix 2**. Draft of the data extraction instrument.

**Appendix 1.**
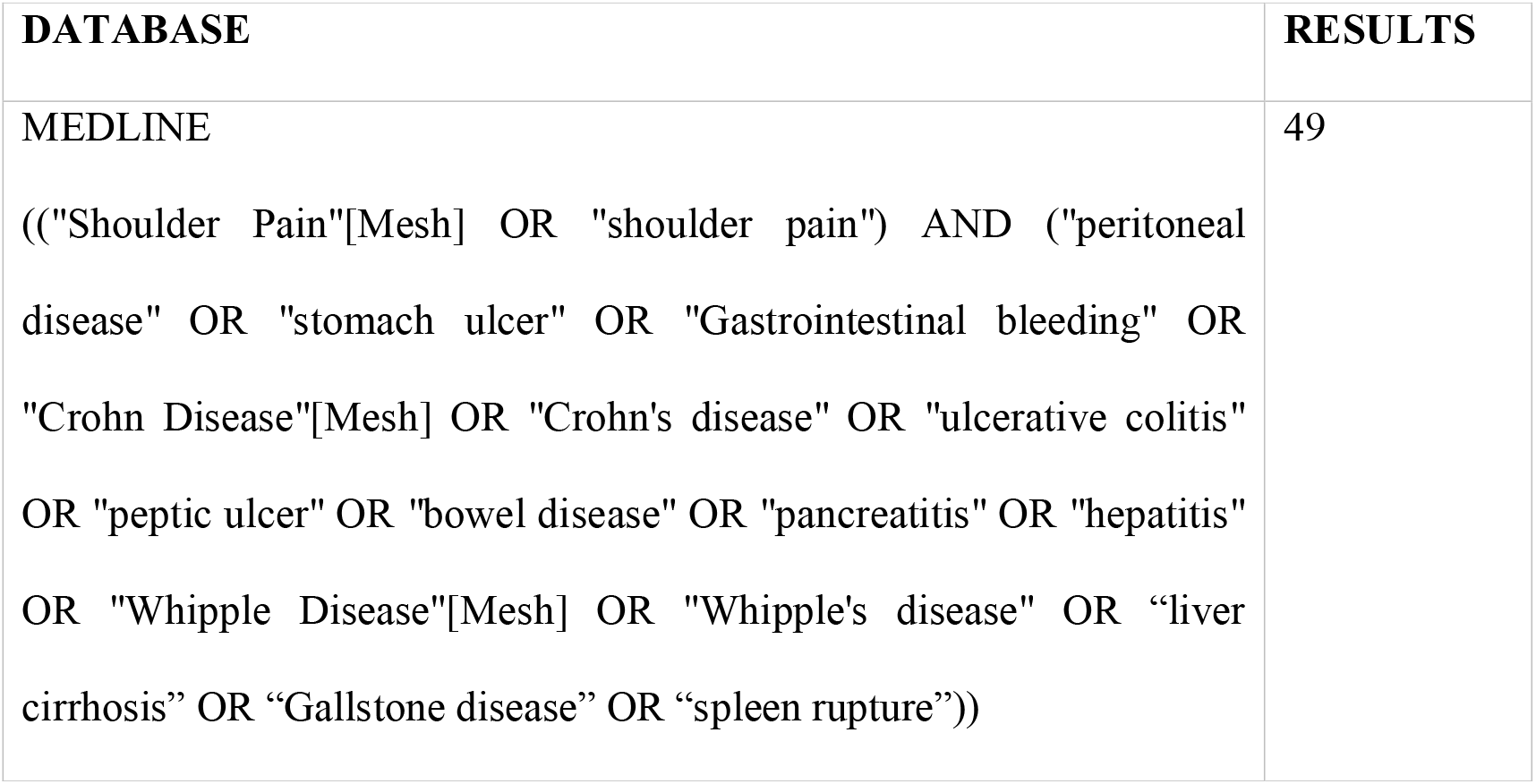
Search conducted on 21^st^ of February 2021.

| DATABASE | RESULTS |
| --- | --- |
| MEDLINE<br>(("Shoulder Pain"[Mesh] OR "shoulder pain") AND ("peritoneal disease" OR "stomach ulcer" OR "Gastrointestinal bleeding" OR "Crohn Disease"[Mesh] OR "Crohn's disease" OR "ulcerative colitis" OR "peptic ulcer" OR "bowel disease" OR "pancreatitis" OR "hepatitis" OR "Whipple Disease"[Mesh] OR "Whipple's disease" OR "liver cirrhosis" OR "Gallstone disease" OR "spleen rupture")) | 49 |

**Appendix 2.**

| Study details |  |
| --- | --- |
| Author(s) |  |
| Year of publication |  |
| Study design |  |
| Population |  |
| Characteristics | e.g. gender, age |
| Sample size |  |
| Diagnosis of shoulder pain |  |
| Concept |  |
| RFs identified | Gastrointestinal and hepatic diseases |
| Context |  |
| Country |  |
| Setting | e.g. private/public clinic |

